# Cardiovascular disease and all-cause mortality risk prediction from abdominal CT using deep learning

**DOI:** 10.1101/2021.08.30.21262686

**Authors:** Daniel C. Elton, Andy Chen, Perry J. Pickhardt, Ronald M. Summers

## Abstract

Cardiovascular disease is the number one cause of mortality worldwide. Risk prediction can help incentivize lifestyle changes and inform targeted preventative treatment. In this work we explore utilizing a convolutional neural network (CNN) to predict cardiovascular disease risk from abdominal CT scans taken for routine CT colonography in otherwise healthy patients aged 50-65. We find that adding a variational autoencoder (VAE) to the CNN classifier improves its accuracy for five year survival prediction (AUC 0.787 vs. 0.768). In four-fold cross validation we obtain an average AUC of 0.787 for predicting five year survival and an AUC of 0.767 for predicting cardiovascular disease. For five year survival prediction our model is significantly better than the Framingham Risk Score (AUC 0.688) and of nearly equivalent performance to method demonstrated in Pickhardt et al. (AUC 0.789) which utilized a combination of five CT derived biomarkers.

## 1. PURPOSE

Globally, cardiovascular disease (CVD) remains the number one cause of mortality, causing 17.9 million deaths in 2019, including 38% of all premature deaths from noncommunicable diseases.^1^ As standards of living increase globally, people have more time and resources to invest in preventative measures to protect their long term health. Since the 1960s, the rate of deaths from CVD in the United States has been reduced by about 50% through a mixture of lifestyle changes and improved treatment. It has been argued that up to 90% of CVD is preventable if lifestyle changes and other interventions are implemented early enough.^2^ Informing a patient that they have higher than average CVD risk can help encourage positive lifestyle changes. Additionally, several primary prevention treatments are under investigation to reduce CVD risk including metformin,^3, 4^ low-dose aspirin,^5^ and gene therapy.^6^ Given the costs and risks associated with these treatments, targeting them to the most at-risk patients will be important.

Genetic screening can help with targeting but completely neglects the effects of environmental exposures. A popular risk scoring system for CVD risk used currently is the Framingham risk score (FRS) which factors in age, sex, cholesterol level, and blood pressure.^7^ In the Framingham Heart Study 8,491 patients were followed for 12 years and it was found that the FRS achieved a C-statistic (equivalent to AUC for censored data) for predicting CVD of 0.763 (95% confidence interval (CI) 0.746 - 0.780) in men and 0.793 (95% CI, 0.772 - 0.814) in women.^8^ Previously we showed that the FRS yields an AUC of 0.688 (95% CI 0.650–0.727) for predicting five year survival for the patient cohort we will study.^9^ A 2019 meta-analysis compared three popular risk models for predicting 10-year CVD risk - the FRS, Framingham Adult Treatment Panel III model, and pooled cohort model.^10^ They found modest C-statistics ranging between 0.68-0.74.^10^

Imaging biomarkers hold much promise for improving CVD risk scoring. In particular, coronary artery calcification (CAC) scoring has been heavily studied.^11, 12^ CAC scores are typically obtained using a specialized ECG gated non-contrast cardiac CT scan. The addition of CAC score has been shown to improve risk prediction when added to more traditional risk factors such as age, gender, blood biomarkers, and family history.^**?**, 13, 14^ In the Multi-Ethnic Study of Atherosclerosis (MESA) dataset the FRS yielded a C-statistic for CVD risk (median follow up time 7.6 years) of 0.62 but that rose to 0.78 when CAC score was included.^15^ Work by Mets et al. obtained a C-statistic of 0.71 (95% CI 0.67-0.76) for three year survival prediction using the National Lung Screening Trial dataset with a multivariate model that combined CAC score and clinical parameters (age, smoking status, etc.)^13^ McClelland et al. obtained a C-statistic of 0.80 for 10 year prediction of coronary heart disease when combining CAC score with blood biomarkers, family history, age, and gender.^14^ Recently several works have shown how deep learning systems can automate CAC scoring.^16–20^ Interestingly, Chao et al. have demonstrated that feeding a box around the heart from a chest CT into a multi-view convolutional neural net (CNN) architecture can lead to improved CVD and mortality prediction over the state of the art for automated CAC scoring or manual CAC grading.^20^ This suggests that CNNs can extract additional features beyond the coronary artery calcification which help inform the risk prediction. Partially by inspired that work, in this work we study feeding an entire abdominal CT scan into a CNN to perform risk scoring rather than extracting custom biomarkers such as aortic calcification score and visceral/subcutaneous fat ratio like we did in a prior work.^9^

The vast majority of prior work on CVD or all-cause mortality risk prediction has looked at either chest CT^19–24^ or chest X-ray.^25, 26^ Among the works that utilize chest CT, the publicly available National Lung Screening Trial (NLST) dataset has been heavily utilized^13, 19, 20, 23, 24, 26^ in addition to the Dutch-Belgian lung cancer screening trial,^21^ or in-house chest CT datasets.^22^ NLST participants have a history of smoking, putting them at higher than average risk for CVD. In contrast, our dataset consists of scans from an otherwise healthy patient cohort. In this work we focus on risk scoring from abdominal CT scans, which has only been studied previously in a handful of works.^9, 27, 28^ Abdominal CT are rich with biomarkers known to be relevant to cardiovascular risk such as visceral fat and aortic plaque.^29, 30^ This is the first work to feed the entire abdominal CT scan into a deep learning model.

## 2. METHODS

The dataset we utilize consists of CT colonography (CTC) scans from the University of Wisconsin Medical Center.^27^ 9,223 people underwent CTC scans between April 2004 and December 2016. Further details on the cohort are provided in Pickhardt et al, 2020.^9^ As in prior works^9, 27^ cardiovascular disease was defined as myocardial infarction, cerebrovascular accident, or development of congestive heart failure. These reflect the endpoints considered by the FRS for cardiovascular disease.

We found 6775 patients had five year follow up data indicating if they survived for five years and out of those 216 (3%) died within five years. Similarly, 7008 patients had follow up data indicating whether or not they were diagnosed with CVD within five years, and out of those 399 developed CVD (5.6%). We used roughly 71% of the data for training, 5% for validation, and 25% for test. The data flow and number of patients in the training, validation, and testing folds for 5 year survival prediction is summarized in figure 2.

The architectures for the CNN and VAE are shown in figure 1. The two architectures were identical apart from the presence of the VAE decoder. To reduce the number of parameters in the network the first convolutional layer contains a large 7×7×7 kernel applied with a stride of 2 in each direction and padding of 3 on the edges. The rest of the convolutional layers use 4×4×4 kernels with a stride of 2 and padding of 1. Group normalization and dropout (dropout rate 0.5) were used after each convolutional layer. In summary, the CNN encoder contains 5 convolutional layers and two feedforward networks and has a total of 5,414,558 parameters. The VAE architecture contains 3 additional fully connected layers and 5 additional upsampling layers and has a total of 21,060,930 parameters. We use a relatively small latent vector size of *D* = 256 to encourage the network to find efficient high level features.

**Figure 1.**
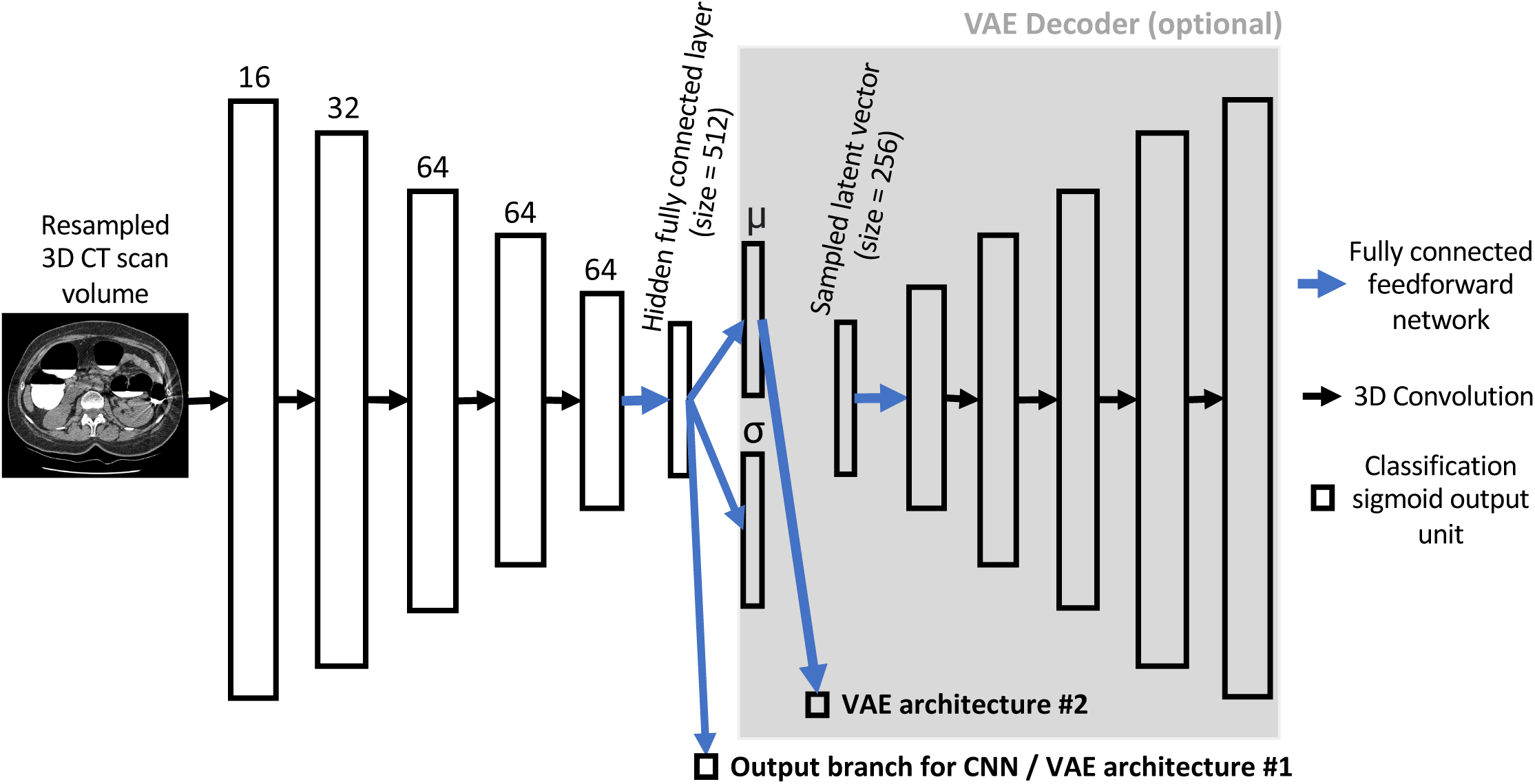
The architectures employed. The CNN architecture consists of five CNN layers followed by two fully connected layers. Optionally a VAE decoder may be attached to the CNN to encourage the CNN to obtain good high level features. If the VAE is attached, one can experiment with attaching the classification output to the vector of latent means (architecture 2)

**Figure 2.**
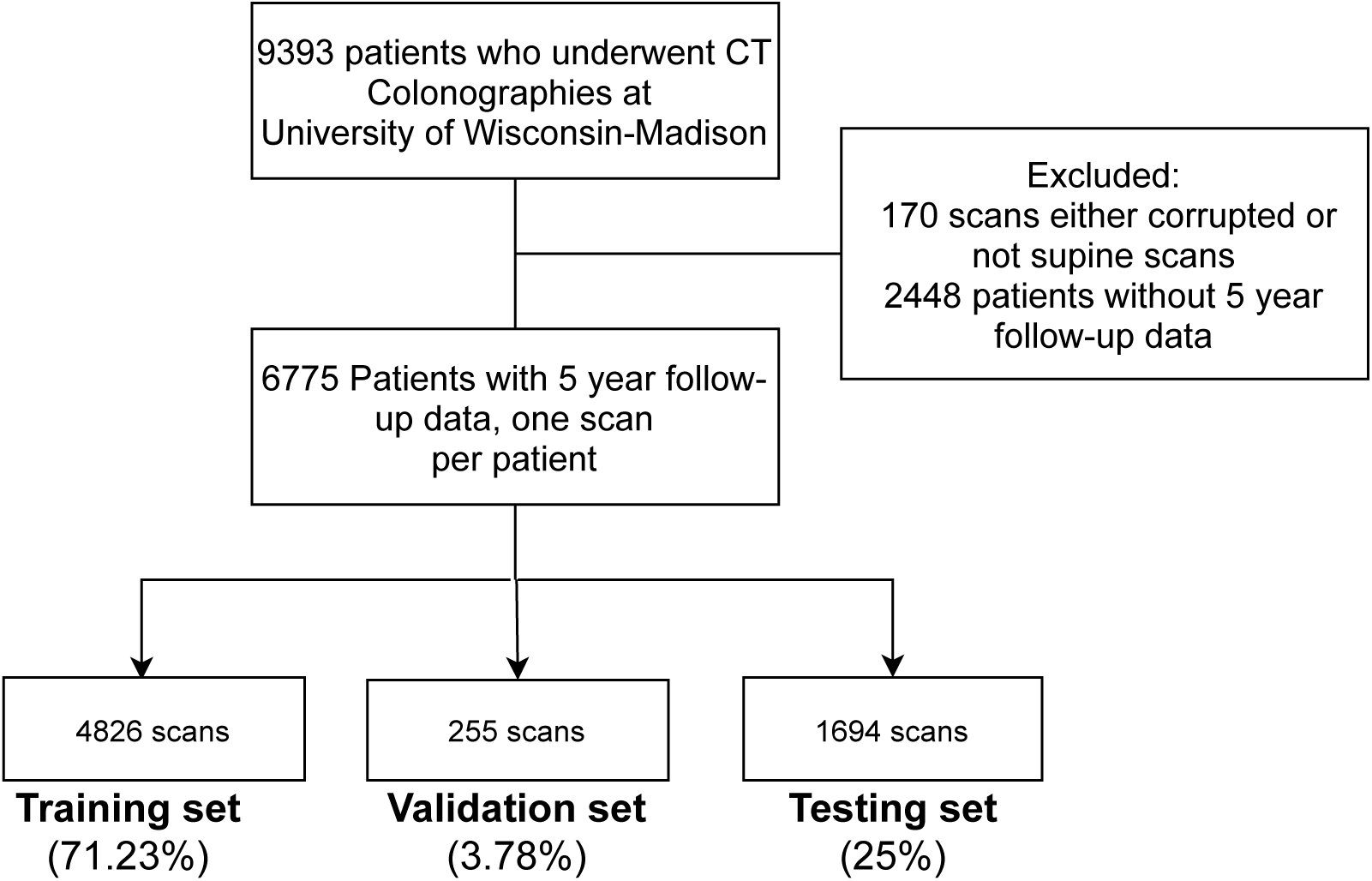
Patient flow chart showing how the patient cohort was selected and split into train, validation, and test sets for five year survival prediction.

We also attempt to add the patient age and gender information to the model. As is standard practice in machine learning, we encode the age and gender information in one-hot vectors. Ages were encoded in a vector of length 60 corresponding to ages between 30-90 years old (any patient with age less than 30 is set to 30 and any above 90 is set to 90). The one hot vectors are concatenated to the hidden layer.

The images are resampled to a size of 192×192×128 using trilinear interpolation, clipped to [-500, 500], and then rescaled to between [0, 1]. To deal with the data imbalance (only ≈ 2% of patients died with 5 years) we reweight the data during training so the ratio of survived/died is 50/50 during training. The RAdam optimizer^31^ was used with a learning rate of 0.001 and batch size of 4. The data augmentations employed were limited to random rotations around a random axis by +/- 0-15 degrees, random 0-20% cropping in the cranial-caudal direction, and random left-right flipping.

Most existing works using a VAE, such as van Velzen et al.,^23^ train an autoencoder first for image reconstruction and then train a separate classification model that utilizes either latent vector or a hidden layer vector taken from the “bottleneck” portion of the VAE. In this work we perform joint training, training our VAE model for both image reconstruction and classification at the same time. This procedure is inspired by previous work where a VAE decoder was attached to a 3D V-Net.^32^ The author claims that adding the VAE helps regularize the 3D V-Net and prevent overfitting.^32^ In a similar way we hypothesize that joint training with the VAE attached will improve the generalization performance of the classifier by encouraging the network to learn robust high level representations. Joint training is more challenging to implement since it requires carefully weighting the loss terms, otherwise one or more loss terms may be ignored during gradient descent training.

The loss function applied at iteration *i* for the VAE architecture was:

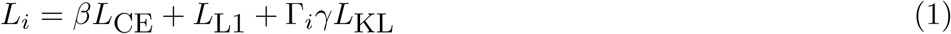

where

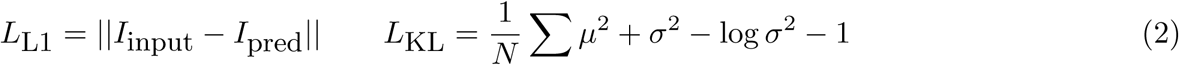

The loss function for the classifier was the standard cross-entropy loss weighted by a factor *β*. For the VAE reconstruction loss we used an L1 loss function. As has been adopted in prior works,^33^ a weighting factor Γ_i_ was applied to Kullback-Leibler loss term of the VAE and linearly increased from 0 to 1 over the course of the first 5,000 iterations. This practice prevents the optimizer from only focusing on minimizing the KL loss in the early stages of training. The factors *β* and *γ* had to be determined empirically. After experimenting with weightings of *β* ∈ {1, 10, 100} and *γ* ∈ {0.001, 0.0001} we used *β* = 10 and *γ* = 0.001 as this was the only combination where all three loss functions decreased during training. Further tuning of these hyperparameters was not attempted. The F1 score and AUC in the validation set were monitored during training. All models were trained for exactly four epochs, which was found to be sufficient for the validation F1 and AUC to converge.

## 3. RESULTS

A summary of the average AUCs obtained in four-fold cross validation is provided in table 3 and select ROC curves are shown in figure 3. In line with our hypothesis, we found that the VAE obtained a higher AUC than the CNN only (0.787 vs 0.768), although we note that the limited number of folds employed means the result is not statistically significant (Welch’s *t*-test *p* = 0.34). VAE architecture option #2, where the classifier is attached to the latent vector means, performed slightly worse than VAE architecture option #1. The concatenation of age and sex information to the final layer of the classifier did not improve the AUC. This may partially be due to the fact that the ages of the patients did not vary that much. It may be possible to better incorporate the age and gender information, eg. by adding an additional fully connected layer.

**Figure 3.**
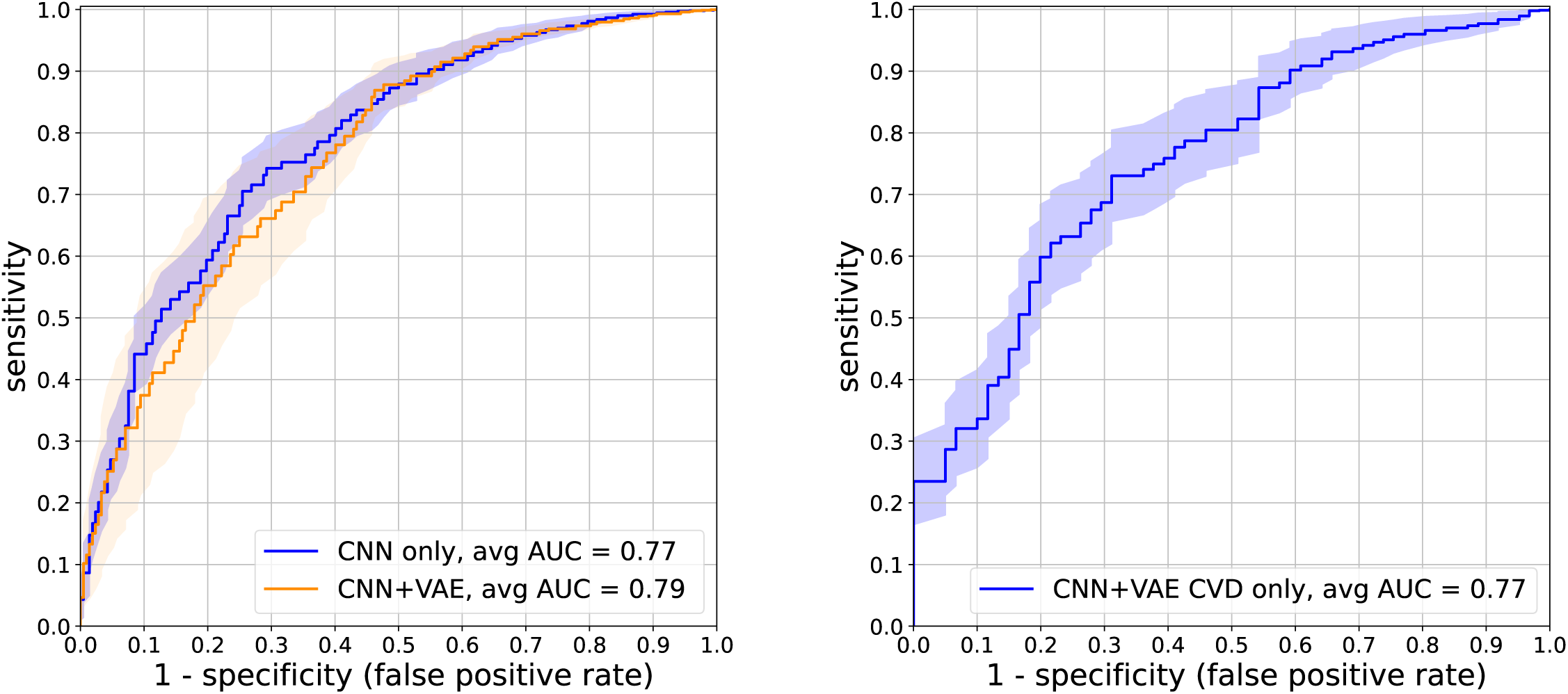
Select average ROC curves under 4-fold cross validation for five year survival prediction (left) and five year CVD prediction (right). The shaded regions show the standard deviation over the four folds.

The results in table 3 were obtained using a box size of 192×192×128. After obtaining those results we ran some hyperparameter optimization experiments to investigate the effect of box size, the results of which are shown in table 3). The results show a slightly lower AUC is found when using a smaller box of size 128×128×128 and a slightly higher AUC is obtained when the box size is increased to 192×192×192, at least for the one fold under consideration. The results suggest that a larger box of 192×192×192 may be beneficial but there is little benefit to increasing the box size beyond that.

## 4. CONCLUSION / NEW OR BREAKTHROUGH WORK

For the first time we have shown how a deep learning model can predict all-cause mortality risk directly from an abdominal CT using a novel jointly trained VAE architecture. We found that the use of the VAE decoder improves the model over using just a plain CNN architecture. The VAE based model is significantly better than using the Framingham Risk Score at predicting five year mortality and is of equivalent accuracy to the best system for abdominal CT demonstrated in Pickhardt et al.^9^ (AUC 0.787 vs 0.789). Our model has the advantage of being much simpler than the prior system, which requires the use of five independent software tools for plaque, bone mineral density, visceral/subcutaneous fat, liver fat, and muscle quantification. On a modern workstation with a NVIDIA Quadro RTX 8000 GPU our model runs in about four seconds and that time reduces to less than one second when our model is preloaded into GPU memory. Many avenues are open for improving our model further.

## Data Availability

The summary numerical output of the automated CT-based tools will be shared upon request, subject to an internal review by PJP and RMS to ensure that participant privacy is protected and subject to completion of a data sharing agreement, as well as approval from both the University of Wisconsin School of Medicine & Public Health (Madison, WI, USA) and the US National Institutes of Health Clinical Center (Bethesda, MD, USA). Pending the aforementioned approvals, data sharing will be made in a secure setting, on a per-case-specific manner. Please submit such requests to PJP.

## ACKNOWLEDGMENTS

This research was supported in part by the Intramural Research Program of the National Institutes of Health Clinical Center.

---

**Table 1.** Summary of average AUCs in four-fold cross validation and standard deviation over the four folds. “CVD+mortality” refers to risk prediction for either CVD diagnosis or death.

| method | AUC 5-year mortality | AUC 5-year CVD or mortality | AUC 5-year CVD |
| --- | --- | --- | --- |
| CNN only | 0.768±0.026 |  |  |
| CNN+VAE (arch #1) | 0.787±0.030 |  | 0.767 ± 0.036 |
| CNN+VAE (arch #1) + age+sex | 0.777±0.031 |  |  |
| CNN+VAE (arch #2) | 0.770±0.034 |  |  |
| FRS<sup>9</sup> | 0.688 | 0.695 |  |
| BMI<sup>9</sup> | 0.499 | 0.552 |  |
| best CT derived model from Pickhardt et al. 2020<sup>9</sup> | 0.789 | 0.742 |  |
| best multivariate model (CT biomarkers+FRS)<sup>9</sup> | 0.796 | 0.751 |  |

**Table 2.** Hyperparameter optimization experiments testing different box sizes for five year survival prediction using the CNN-only architecture. Validation AUC is an average of the validation AUCs obtained in the the last epoch of training and the test AUC is the AUC on the hold-out test set for the final model after 4 epochs of training.

| box size | validation AUC | test AUC |
| --- | --- | --- |
| 128x128x128 | 0.75 | 0.74 |
| 192x192x128 | 0.76 | 0.76 |
| 192x192x192 | 0.78 | 0.79 |
| 256x256x128 | 0.78 | 0.76 |

